# Impact of COVID-19 related healthcare changes on antibiotic resistance in clinical *Escherichia coli* isolates: interrupted time series analyses in Scotland, UK

**DOI:** 10.1101/2025.05.22.25328070

**Authors:** Laura Ciaccio, Peter T Donnan, Benjamin J Parcell, Charis A Marwick

**Affiliations:** Division of Population Health and Genomics, School of Medicine, University of Dundee, Dundee

**Author notes:** Corresponding Author: Laura Ciaccio, University of Dundee, School of Medicine Mailbox 1, Level 7, Corridor L Ninewells Hospital Dundee, DD1 9SY.

## Abstract

**Objectives:** The SARS-CoV-2 pandemic has impacted healthcare use, with mixed reports about the impact on antimicrobial resistance. This work aimed to identify changes in healthcare utilisation and antibiotic prescribing in relation to the COVID-19 pandemic and quantify any subsequent impact on antibiotic resistance in clinical *Escherichia coli* isolates across a complete geographical population in Scotland.

**Methods:** Data including ∼490,000 people from January 2018 to March 2022 were accessed via the University of Dundee. Joinpoint regression analyses identified changes in trend for hospital encounters and antibiotic use in the community and hospital. Using identified joinpoints as the “intervention” time point, the impact of these changes on the proportions of *E. coli* blood and urine culture isolates that were antibiotic resistant and multidrug resistant (MDR) were examined using interrupted time series analysis (ITSA).

**Results:** Joinpoint regression analyses identified January 2020 as the intervention time point for ITSA. From 26% resistant (not MDR) and 35% MDR among urine *E. coli* isolates in the month immediately pre-intervention, there were upward changes in level of 2.5% (95%CI - 0.4% to 5.4%) and trend of 0.3% (95%CI 0.1% to 0.5%) per month for resistant (not MDR), and an upward change in level of 0.4% (95%CI - 2.0% to 2.8%) and a downwards change in trend of −0.3% (95% CI - 0.5% to −0.1%) per month for MDR. Increases in resistant (not MDR) and reductions in MDR proportions were found 9 months post-intervention compared to proportions predicted had the intervention not occurred. Similar changes were observed for blood culture isolates, but numbers were smaller, resulting in less certainty around estimates.

**Conclusion:** There were small but significant reductions in the proportions of MDR *E. coli* isolates associated with COVID-19-related changes to healthcare utilisation and antibiotic prescribing. These results may inform future antimicrobial stewardship practices and their evaluation, including estimates of their impact on antibiotic resistance.

## Introduction

The World Health Organization (WHO) declared COVID-19, caused by the SARS-CoV-2 virus, a pandemic on the 11th of March 2020. Healthcare delivery was severely impacted by measures to contain the spread of COVID-19, such as UK lockdown restrictions, international travel restrictions, modifications to infection prevention and control (IPC) measures, COVID testing, vaccination delivery initiatives, and also by surges of severely ill patients during pandemic waves.^1,2^

These changes in healthcare could plausibly impact outcomes not directly related to COVID-19 infections, such as antibiotic resistance among bacterial infections. Antibiotic resistance is a global public health threat,^3^ associated with prolonged illness, increased risk of transmission and disease, healthcare costs, and mortality.^3^ Understanding the impact of the changes in healthcare utilisation and antibiotic prescribing related to the COVID-19 pandemic could help inform prevention of future antibiotic resistance. Previous studies of antibiotic resistance during the COVID-19 pandemic mainly focus on hospital inpatients with COVID-19,^4–7^ but contributing factors and impacts on resistance are likely to be much broader, and most study designs do not provide measurable effects over time in the same population.

One study using segmented regression analyses reported small reductions in resistance to five, and a small increase to one, antibiotics in *Escherichia coli* blood culture isolates related to COVID-19 restrictions,^8^ but did not account for changes in antibiotic use or resistance in other infections.^9^

This work aims to identify the timing of the impact of the SARS-CoV-2 pandemic on healthcare utilisation and antibiotic prescribing and potential downstream effects on antibiotic resistance in *E. coli* in clinical isolates in a complete geographic region of Scotland, U.K.

## Methods

### Data sets and access

Anonymised data were made available by the Health Informatics Centre (HIC) at the University of Dundee within an established trusted research environment (TRE). The study cohort included everyone registered with a general practice in the National Health Service (NHS) region of Tayside in 2018 and/or 2020.

Datasets included hospital admissions, accident and emergency, general practice out-of-hours, NHS24 (telephone triage service), Scottish Ambulance Service (SAS), community antibiotic prescriptions, hospital antibiotic use, and microbiology.^10^ All data were examined as monthly counts or rates. Encounters with different acute health services were linked into “episodes of care” if they occurred within 24 hours of each other and/or had common episode identifiers. Antibiotic use was measured as dispensed prescribed items (“prescriptions”) in the community and as defined daily doses (DDD)^11^ dispensed at the ward level in hospitals. Antibiotics with potential action against Gram-negative bacteria (see web-only Supplementary Table S1), including *E. coli,* were included in the analysis.

Numbers of *E. coli* blood and urine isolates were examined (see web-only Supplementary Figures S1 and S2). *E. coli* isolates were categorised as susceptible or resistant using results of routine NHS laboratory testing against a standard panel of antibiotics, which uses Vitek 2 (bioMérieux, Marcy-l’Étoile, France) and the European Committee on Antimicrobial Susceptibility Testing (EUCAST) criteria for minimum inhibitory concentration (MIC) cut-off values.^12^ Resistant isolates were further categorised into multidrug resistant (MDR), defined as resistant to at least one or more antibiotics from three or more antibiotic categories (adapted from an international consensus definition^13^) and resistant (not MDR) (see web-only Supplementary Table S2).

### Joinpoint Regression Analysis

Joinpoint regression analyses were used to examine changes in trends in healthcare and antibiotic use. Joinpoint software identifies any point of trend change, or “joinpoint”, and calculates the rate of change and statistical significance,^14^ without user-specification of time points of interest (see methods supplement for additional detail).^15^ Joinpoints plausibly attributable to the COVID-19 pandemic (i.e., from December 2019 forward) were considered for inclusion as the interruption (or “intervention”) timepoint for interrupted time series analysis (ITSA) analysis of the impact on antibiotic resistance. The command line version of the Joinpoint Regression Program 4.9.1.0 was obtained from the National Cancer Institute (NCI).^15^

### Interrupted Time Series Analysis (ITSA)

ITSA used linear models (generalised least squares fit by maximum likelihood), adjusted for autocorrelation, to examine changes in antibiotic resistance among *E. coli* blood and urine culture isolates associated with identified joinpoint(s). The outcomes were proportions of isolates per month that were resistant (not MDR) and MDR.

Summary model outputs included the intercept (β_0_), the pre-intervention trend (β_1_), and changes in level (β_2_) and trend (β_3_) at the intervention point, with 95% confidence intervals (95%CI) and p-values. Post-intervention trends were calculated (β_1_ + β_3_) with 95%CI (see methods supplement).

Differences between observed proportions of resistant (not MDR) and MDR isolates following the intervention and proportions that might have been observed had the intervention not occurred were estimated two ways. First, proportions based on modelled actual values at 3-, 6-, and 9-months post-intervention were compared to values from counterfactual models assuming the pre-intervention trend continued and, second, to conservative models assuming the trend levelled off at the intervention point. Calculation of 95% confidence intervals around the estimated differences while incorporating uncertainties around multiple estimates, produces 95%CI that quickly become too wide to be interpretable. These were not calculated or reported, and estimates should be considered illustrative and not statistically significant. All analyses used RStudio version 4.1.2.

## Results

The study population included 490,021 individuals, with 577,536 episodes of acute care involving 271,208 individuals in the time series. There were five joinpoints identified, in January 2020, April 2020, September 2020, February 2021, and June 2021 (Figure 1A). Prior to the January 2020 joinpoint, healthcare episodes were increasing by 131.5 (95%CI 105 to 158) episodes per month. There was a trend change at the joinpoint of −1,485.7 (95%CI - 2,763 to −208) per month, resulting in a downward trend of −1,354 (95%CI −2,631 to −77.0) episodes per month (January to April 2020).

**Figure 1.**
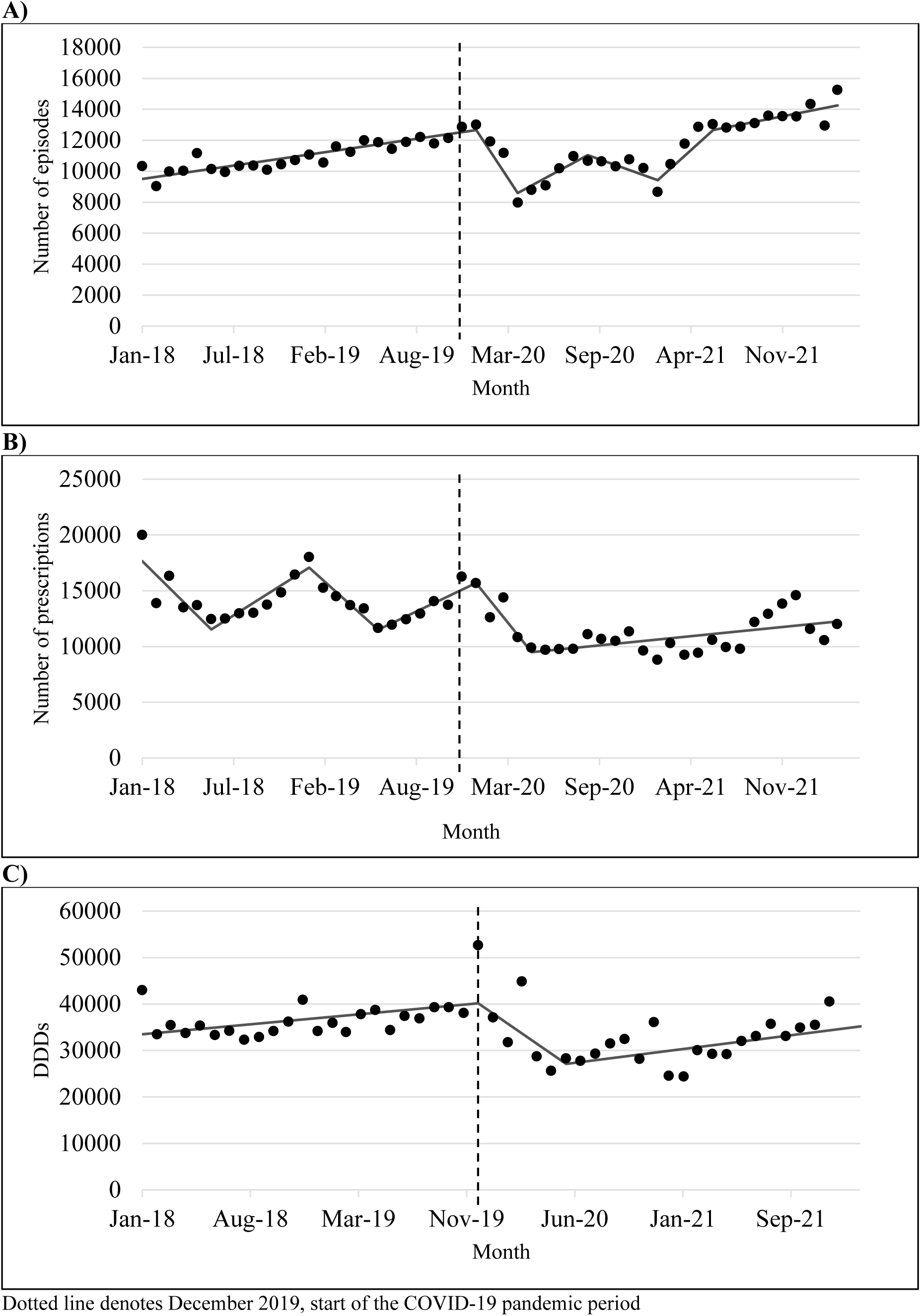
Trends with joinpoints for A) episodes of care, B) community prescribing of antibiotics with action against Gram-negatives, and C) hospital dispensing of antibiotics with action against Gram-negatives

There were 642,716 prescriptions for relevant antibiotics (see web-only Supplementary Table S1) in the community from January 2018 to March 2022. Five joinpoints were identified (Figure 1B), with both joinpoints in the COVID-19 period aligning with joinpoints for healthcare episodes, including January 2020. Prior to the January 2020 joinpoint, prescriptions were increasing by 537.5 (95%CI −62.9 to 1,138) prescriptions per month, with a change at the joinpoint of −2,019.4 (95%CI −3,894 to −145) per month resulting in a downward trend of −1,481.9 (95%CI −3,258 to 294) prescriptions per month (January to March 2020).

There were 1,646,791 DDD of relevant antibiotics dispensed to hospital wards for inpatient use from January 2018 to December 2021. Two joinpoints were identified in December 2019 and May 2020 (Figure 1C). Prior to the December 2019 joinpoint, dispensing of relevant antibiotics was increasing by 288 (95%CI 70.0 to 507) defined daily doses (DDD) per month with a −2,463 (95%CI −4,672 to −254) change at the joinpoint resulting in a downward trend of −2,175 DDD (95%CI 4,372 to 23.1) per month after the joinpoint.

January 2020 was selected as the intervention time point for ITSA based on healthcare and community antibiotic use, incorporating changes following the December 2019 change in hospital antibiotic use.

### ITSA of antibiotic resistance in *E. coli* isolates

There was a total of 32,467 patients (6.6% of the population) with an *E. coli* blood and/or urine isolate during the study period. There were 58,410 *E. coli* urine isolates involving 29,384 patients. The number of *E. coli* urine isolates per month reduced in early 2020 at the start of the pandemic, though this was in line with the reductions seen in the overall number of urine specimen at this time (see web-only Supplementary Figure S1).

In ITSA analysis of urine isolates, there was a statistically significant downward pre-intervention trend in the proportion of *E. coli* that were resistant (not MDR) of −0.2% per month (Table 1, Figure 2A). At the intervention point, there were upward changes in level of 2.5% (95%CI −0.4% to 5.4%) and trend of 0.3% (95%CI 0.1% to 0.5%) per month, with an upward post-intervention trend of 0.1% (−0.1% to 0.3%). For MDR, there was a slight upward pre-intervention trend of 0.05% (95%CI −0.1% to 0.2%) per month and a small upward change in level of 0.4% (95%CI −2.0% to 2.8%) at the intervention. There was a change in trend of −0.3% (95% CI −0.5% to −0.1%) in January 2020, resulting in a significant downward post-intervention trend of −0.2% (95%CI −0.4% to −0.02%) per month (Table 1, Figure 2B).

**Figure 2.**
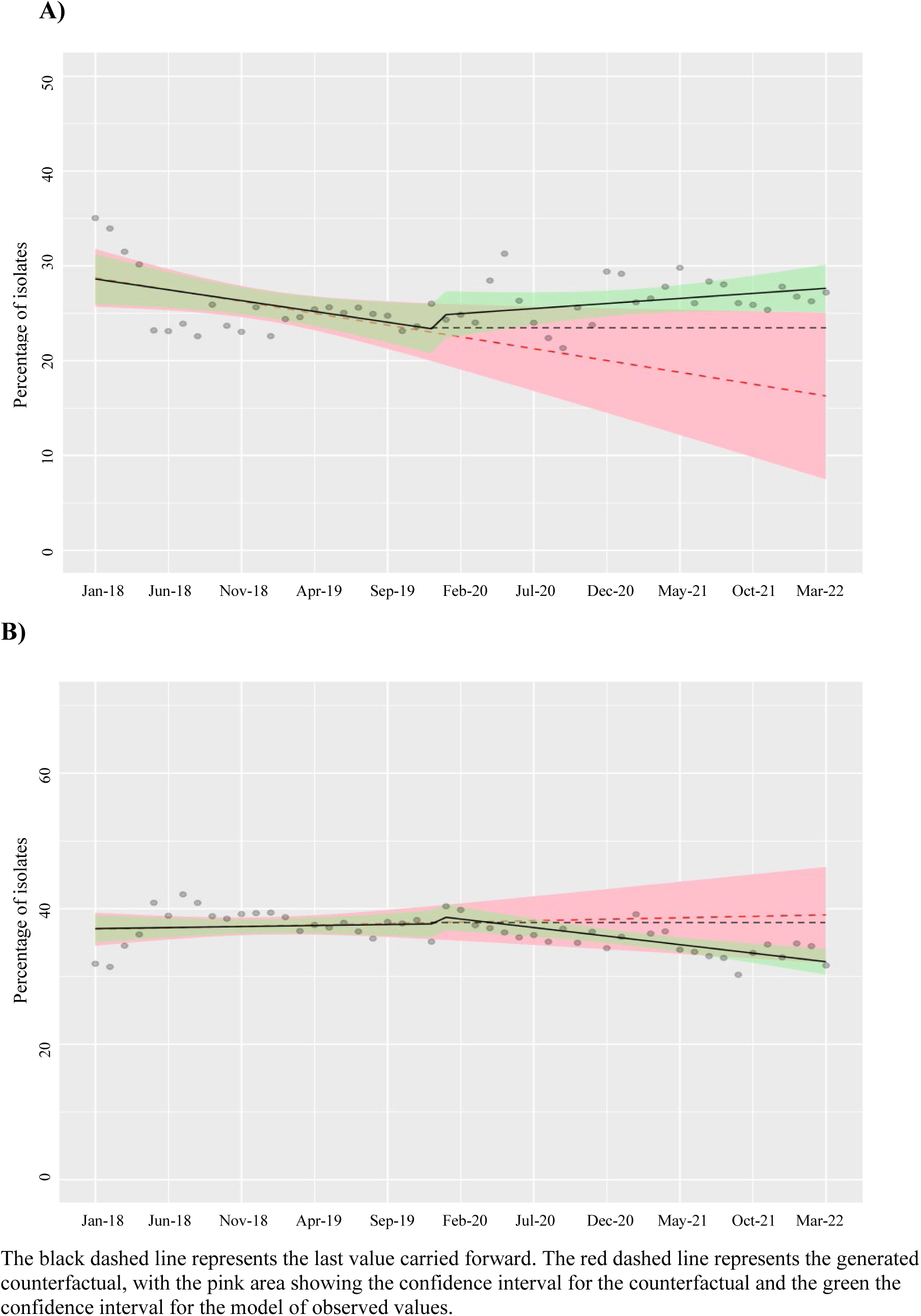
Interrupted time series analysis of the percentage of A) resistant (not-MDR) and B) MDR urine isolates

**Table 1.** Level and trend changes in the proportions of resistance in urine isolates associated with January 2020 intervention from interrupted time series analysis.

| Parameters in Model | Coefficient<br>(by Month) | 95% Confidence<br>Interval | P-Value |
| --- | --- | --- | --- |
| Resistant (not MDR) |  |  |  |
| Intercept ($\beta_0$) | 28.6% | 26.6% to 30.6% | <0.001* |
| Pre-Intervention Trend ($\beta_1$) | -0.2% | -0.4% to -0.1% | 0.002* |
| Level Change, January 2020 ($\beta_2$) | 2.5% | -0.4% to 5.4% | 0.09 |
| Trend Change, January 2020 ($\beta_3$) | 0.3% | 0.1% to 0.5% | 0.003* |
| Post-Intervention Trend ($\beta_1+\beta_3$) | 0.1% | -0.1% to 0.3% | - |
| MDR |  |  |  |
| Intercept ($\beta_0$) | 37.1% | 35.4% to 38.7% | <0.001* |
| Pre-Intervention Trend ($\beta_1$) | 0.05% | -0.1% to 0.2% | 0.47 |
| Level Change, January 2020 ($\beta_2$) | 0.4% | -2.0% to 2.8% | 0.74 |
| Trend Change, January 2020 ($\beta_3$) | -0.3% | -0.5% to -0.1% | 0.002* |
| Post-Intervention Trend ($\beta_1+\beta_3$) | -0.2% | -0.4% to -0.02% | - |

At nine months post-intervention, the proportion of resistant (not MDR) urine isolates was estimated at 5.2% higher than counterfactual (20.6% vs 25.8%), and 2.5% higher than conservative estimates. For MDR, proportions were estimated at −1.9% (38.4% vs 36.5%) and −1.3% lower than predicted, respectively (see web-only Supplementary Table S3).

There were 1,683 *E. coli* blood culture isolates identified for 1,344 patients. The relatively low number of blood cultures and subsequently low number of blood culture isolates (15 to 47 per month) results in highly variable trends (see web-only Supplementary Figure S2)

In ITSA analysis, there was a slight downward pre-intervention trend in the proportion of resistant (not MDR) isolates (Table 2, Figure 3A). There was a small downward level change of −1.7% (95%CI −11.3% to 7.9%) and upward trend change of 0.4% (95%CI −0.2% to 1.0%) at the intervention time point, resulting in a post-intervention trend of 0.02% (95%CI −0.8% to 0.9%). For MDR, there was an upward pre-intervention trend of 0.4% (95%CI −0.2% to 0.9%) (Table 2, Figure 3B). There were negative level (−2.8% [95%CI −13.0% to 7.4%]) and trend (−0.4% [95%CI −1.2% to 0.4%]) changes, as well as a downward post-intervention trend of −0.1% (95%CI −1.0% to 0.8%).

**Figure 3.**
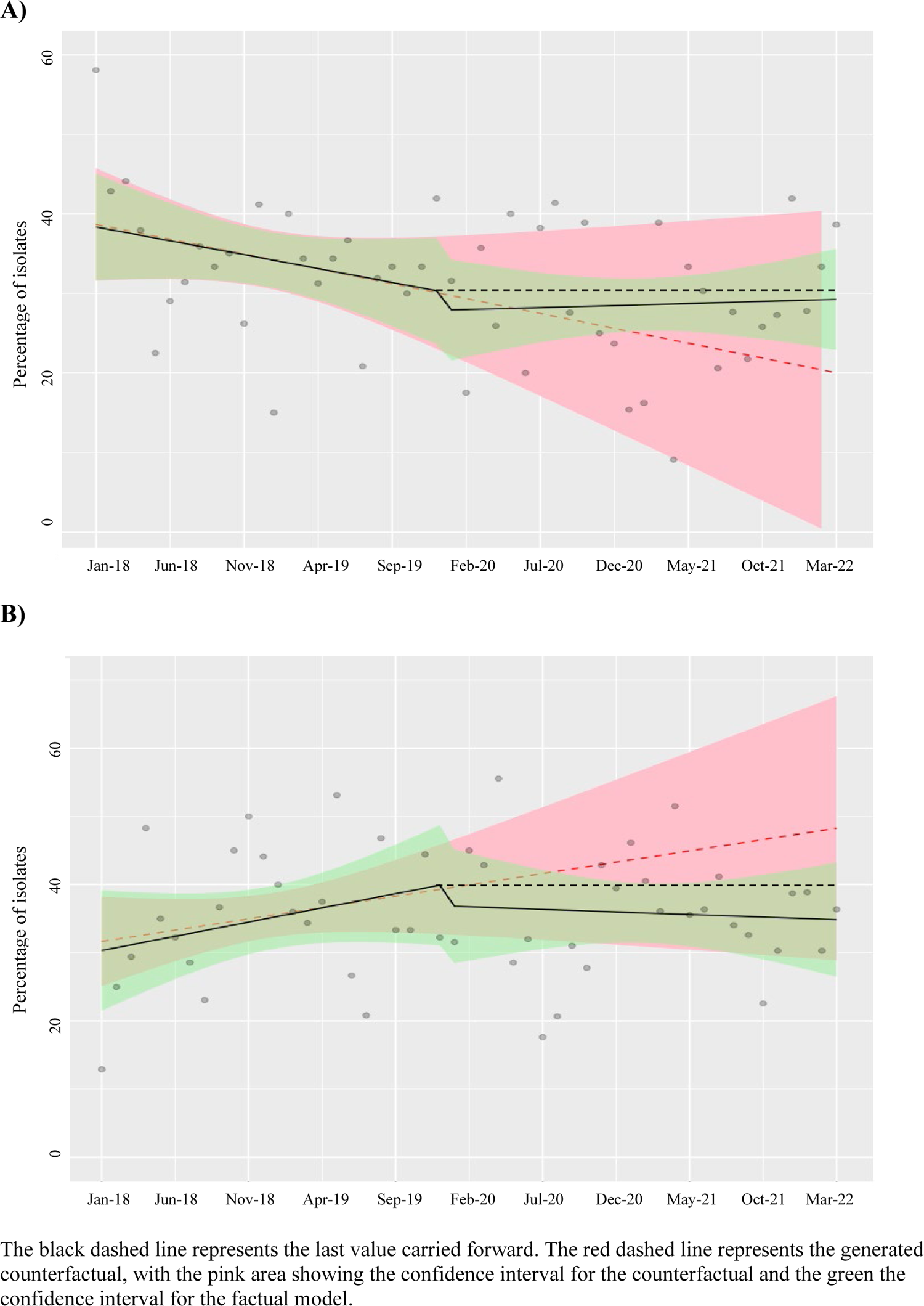
Interrupted time series analysis of the proportion of A) resistant (not-MDR) and B) MDR blood culture isolates

**Table 2.** Level and trend changes in the proportions of resistance in blood culture isolates associated with January 2020 intervention from interrupted time series analysis.

| Parameters in Model | Coefficient<br>(by Month) | 95% Confidence<br>Interval | P-Value |
| --- | --- | --- | --- |
| Resistant (not MDR) |  |  |  |
| Intercept ($\beta_0$) | 38.4% | 31.6% to 45.2% | <0.001 |
| Pre-Intervention Trend ($\beta_1$) | -0.4% | -0.9% to 0.1% | 0.16 |
| Level Change, January 2020 ($\beta_2$) | -1.7% | -11.3% to 7.9% | 0.26 |
| Trend Change, January 2020 ($\beta_3$) | 0.4% | -0.2% to 1.0% | 0.73 |
| Post-Intervention Trend ($\beta_1+\beta_3$) | 0.02% | -0.8% to 0.9% | - |
| MDR |  |  |  |
| Intercept ($\beta_0$) | 31.1% | 23.8% to 38.3% | <0.001 |
| Pre-Intervention Trend ($\beta_1$) | 0.4% | -0.2% to 0.9% | 0.18 |
| Level Change, January 2020 ($\beta_2$) | -2.8% | -13.0% to 7.4% | 0.59 |
| Trend Change, January 2020 ($\beta_3$) | -0.4% | -1.2% to 0.4% | 0.22 |
| Post-Intervention Trend ($\beta_1+\beta_3$) | -0.1% | -1.0% to 0.8% | - |

At nine months post-intervention, the proportion of resistant (not MDR) isolates was estimated to be 2.0% higher than counterfactual, and −2.0% lower than conservative estimates. For MDR, proportions were −6.5% and −3.8% lower than estimates, respectively (see web-only Supplementary Table S4).

## Discussion

This population-level study identified changes in healthcare and antibiotic use attributable to the COVID-19 pandemic and possible downstream impact on antibiotic resistance in clinical *E. coli* isolates. Following January 2020 at the start of the COVID-19 pandemic, there were downward changes in trend for MDR among urine and blood culture isolates, with upward changes in trend for resistant (not MDR).

For urine isolates, the changes in trend were inverse at −0.3% and 0.3% for MDR and resistant (not MDR) isolates, respectively. Similarly, there was also a balance in trend changes seen for the blood culture isolates (0.4% and −0.4%). By October 2020, the proportion of resistant (not MDR) isolates were estimated to have increased between 2.5% (conservative estimate) and 5.2% (counterfactual) compared to the actual modelled values, while the MDR proportion was estimated to decrease between 1.3% (conservative estimate) and 1.9% (counterfactual). Had the previous trends continued, 59.0% of urine isolates would have been resistant overall (including MDR) in October of 2020, with the actual modelled values being 62.3%, which is higher than if the pandemic had not occurred (with the caveat of imprecise estimates).

### Strengths and limitations

This study used large population-level datasets, including routine healthcare data, increasing the generalizability of the results and minimising missing data. In analyses of prescribing practice in Scotland, there were different measures of antibiotic use for community and hospital, with neither able to measure consumption. However, the community prescribing dataset includes items dispensed rather than just prescribed, which takes it closer to capturing consumption.

The study findings may not be generalisable to areas with different demographic characteristics, especially as ethnicity data were unavailable, but the Tayside region is demographically representative of the Scottish population.^16^

### Comparison with other studies

Other studies report varying changes in antibiotic resistance due to the COVID-19 pandemic. A multicentre cross-sectional study in Egypt compared 2019 to 2022 and found a statistically significant increase in the proportion of MDR *E. coli* isolates (64% vs 61%, respectively).^17^ This contrasts our observed decrease, and that reported in other studies,^18–21^ but they used an uncontrolled before-after design which does not account for underlying secular trends. One retrospective study in Indonesia comparing *E. coli* resistance in the first six months of the pandemic with the six months prior demonstrated a decrease in Extended-spectrum beta-lactamase (ESBL)-producing *E. coli* isolates, with an increase in rates of susceptibility to ten of sixteen antibiotics tested.^18^ Another study of *E. coli,* in the Dominican Republic pre- and post-pandemic, reported that the proportion of ESBL-producing *E. coli* decreased from 25.6% to 24.8%.^19^ A systematic review of antibiotic resistance during COVID-19 also reported decreases in resistance for *E. coli*, including a 2.4% decrease (to 58.2%)in ESBL-producing *E. coli* isolates despite an increase in resistance across other bacteria examined.^20^ Another systematic review reported increased Gram-negative resistance during the pandemic, and that *Acinetobacter baumannii* and *Klebsiella. pneumonia* had higher rates of resistance than *E. coli.*^21^ Many of the studies included in both systematic reviews examined ICU patients where rates of resistant organisms are often higher than among the general population.^22–24^ Regarding the overall rates of resistance in our study, which examined a whole geographic population, there was only an overall increase in resistance for urine *E. coli* isolates.

One study which used ITSA to examine changes in resistance among *E. coli* blood culture isolates to six individual antibiotics and antibiotic classes in a geographic region of England from 2016 to 2021.^8^ They reported decreases in resistance to piperacillin-tazobactam, ciprofloxacin, co-amoxiclav, gentamicin, and third-generation cephalosporins following the start of COVID-19 restrictions. Carbapenems were the only antibiotic examined with an increase in resistance during the pandemic, and this was numerically very small.^8^ Carbapenem resistance was exceptionally low in our population, excluding formal analysis. These changes appear broadly similar to our findings, but it is difficult to make a direct comparison as they did not report MDR rates.

National routine surveillance reports state that resistance rates in urine and blood culture *E. coli* isolates in Scotland remained stable overall between 2020 and 2022, with small changes in resistance to individual antibiotics.^25^ There were decreases in resistance in blood culture isolates for co-amoxiclav and piperacillin-tazobactam, and in urine isolates for co-amoxiclav, nitrofurantoin, trimethoprim, and piperacillin-tazobactam.^25^ Temocillin resistance was more variable, and fosfomycin resistance in urine isolates increased.^25^ These decreases in resistance to commonly tested antibiotics align with our observed decrease in MDR, although “MDR” is not reported in national Scottish surveillance data.

### Implications for policy and practice

The results of this work in the context of the COVID-19 pandemic have the potential to help inform future policies and practices during disruptions to healthcare. The early pandemic response holds the greatest potential for learning regarding future pandemic preparedness. The data presented in this work demonstrate the initial impact of the pandemic on the number of episodes of care and antibiotic use during the first pandemic wave. During the early phase of the second pandemic wave in late 2020, even as vaccination programs had not yet been rolled out to the public, the impact on health utilisation was on a much smaller scale than in wave 1. This suggests that practices put in place during the first wave allowed the later impacts on healthcare to be less pronounced, despite the second wave having the largest number of cases of the three waves covered by this work.^26^

Data now available demonstrates that antibiotic use early in the pandemic was higher than necessary based on estimates of low rates of bacterial co-infection.^4,6,27,28^ This emphasises the need for continued antimicrobial stewardship practices, including clear, timely messaging to the public and healthcare professionals during viral pandemics and outbreaks. This work also enables estimates of the effect of antibiotic use on resistance, which may inform future evaluations of the predicted impact of stewardship interventions. The very large changes seen early in the pandemic, which are unlikely to be replicated in response to stewardship activities, resulted in small downstream changes in antibiotic resistance. However, the large changes early in the pandemic were transient, with many rates returning to pre-pandemic levels by the point of assessment of resistance. This emphasises the need for sustained stewardship efforts with resulting large and long-term changes in antibiotic use to make meaningful reductions in antibiotic resistance. It also demonstrates the difficulties in predictive modelling of the impact of changes in prescribing and healthcare use on antibiotic resistance previously reported.^29,30^

## Conclusion

This work has demonstrated the widespread impact of the COVID-19 pandemic on healthcare and resulting small decreases in the proportion of *E. coli* isolates that were MDR, balanced by small increases in the proportion that were antibiotic resistant but not meeting MDR criteria. The magnitude of changes observed highlights that large and sustained changes will be required to significantly impact antibiotic resistance.

## Transparency declaration

The authors have no conflicts of interest to declare.

## Author contributors

Study concept: CAM. Study design: LC, CAM, PTD. Data cleaning: LC. Statistical analysis: LC. Interpretation of data and results: LC, CAM, PTD, BJP. Drafting of the manuscript: LC. Manuscript feedback and revisions: LC, CAM, PTD, BJP. Administrative, technical, or material support: LC, CAM, PTD, BJP. All authors have read and approved the manuscript for submission.

## Data Sharing

Data from this study are not publicly available but may be accessed by approved researchers via application to the Health Informatics Centre at the University of Dundee.

## Ethics statement

Ethics committee review was not required for this study. All work using these data was completed in the Health Informatics Centre (HIC) secure virtual trusted research environment (TRE) with access controlled by HIC for approved users (ISO27001 accredited). HIC standard operating procedures (SOP) are approved by the East of Scotland Research Service and the NHS. Individual studies complying with these SOPs do not require individual ethics review.

