## Supplemental Methods and Materials for "Impact of COVID-19 related healthcare changes on antibiotic resistance in clinical *Escherichia coli* isolates: interrupted time series analyses in Scotland, UK"

**Methods Supplement**

Joinpoint Regression Analyses

To examine changes in healthcare utilisation and antibiotic prescribing, joinpoint regression analyses were used. Joinpoint regression is a statistical method used to analyse trends in data over time, identify any changes in trends, referred to as "joinpoints", and calculate the statistical significance of these changes and the change in slope following a joinpoint.[1] A key feature of joinpoint analysis, compared to other time series analysis methods, is that there is no user-specified time point to evaluate changes in relation to – the software detects and quantifies joinpoints anywhere in the time series. The command line version of the Joinpoint Regression Program was obtained from the National Cancer Institute (NCI), which developed the software.[2]

The joinpoint regression software divides time series data into linear segments on a log scale.[2–4] The algorithm used to identify the joinpoints then looks for a significant change in the trend line slope between segments. For the version of software provided (v4.9.1.0), a maximum of five joinpoints can be identified for any time series.[2] Once a change in slope is identified, the software uses a Monte Carlo Permutation method to test the significance of the difference in the trend line between adjacent segments. The reported results include the identified joinpoints, with 95% CIs for each, the slope of the line for each segment, the intercept for each line segment if it were to be projected back to the start of the time series, and the change in slope following the joinpoint. For the slope and change in slope, a p-value is provided with the significance level set at $<0.05$. A standard error is also provided for each value, allowing for the calculation of the 95% confidence intervals for the slope and slope change (1.96 $\pm$ [2xSE]).

Joinpoint regression was applied to the episodes of care, antibiotics with potential action against Gram-negative prescribed in the community, and antibiotics with potential action against Gram-negative used in hospital. The slope and intercept values were then used to calculate the modelled values at each joinpoint (y=m(x)+b), and trend lines were added between joinpoints to aid visual interpretation (Figure 10). From the joinpoints identified in each dataset, the first change in trend identified that could potentially be attributed to the COVID-19 pandemic (from December 2019 forward) was selected as a change point of interest for analysis of the impact of COVID-19-related changes on antibiotic resistance.

Interrupted Time-Series Analysis (ITSA)

For the ITSA, in contrast to the joinpoint regression analysis, the intervention time point was user-specified for the analysis. In this work, the specified time point was the result of the joinpoint analysis. Segmented regression was used to examine the proportion of isolates that were resistant (not MDR [as defined in main manuscript]) and the proportion that were MDR for blood and urine culture *E. coli* isolates, examined in separate models. Level and trend changes following the designated intervention time point were identified using segmented regression. In addition to the packages mentioned during previous analyses, nlme, ggplot2, and AICcmodavg were used in Rstudio.[5]

Once the datasets were appropriately formatted, a linear model (generalised least squares fit by maximum likelihood) was run on each data set. The summary output from these models included the intercept ($\beta_0$), pre-intervention trend ($\beta_1$), change in level ($\beta_2$) and change in trend ($\beta_3$) at the intervention point, with 95% confidence intervals and p-values for each. The post-intervention trends were calculated ($\beta_1 + \beta_3$), with 95% confidence intervals. The outputs were graphed in R with confidence intervals generated around the modelled trend lines.

After generating the initial model, testing for significant autocorrelation was completed to determine the necessary autoregressive order (p) and moving average order (q) for the projection to improve the Akaike Information Criterion (AIC), which is a measure of goodness-of-fit (lower is better) for each model (see web-only Supplementary Table S5).[5] The p and q values for final models were selected based on the AIC visual best fit to the pre-intervention trend.

A projected, counterfactual model was used to estimate the difference between the modelled values (actual versus predicted by counterfactual) at 3, 6, and 9 months post-intervention had the intervention not occurred. An additional projected model that assumed the trend levelled off at the intervention point was generated in R to provide more conservative predicted values had the intervention not occurred and calculate estimates between these and actual modelled values at 3, 6 and 9 months.

### Supplemental materials

#### Table S1- Relevant antibiotics by approved name included in analysis

| Approved name |
|---|
| Amoxicillin |
| Ampicillin |
| Ampicillin With Cloxacillin |
| Cefaclor |
| Cefalexin |
| Cefixime |
| Cefradine |
| Cefuroxime |
| Chloramphenicol |
| Ciprofloxacin |
| Co-amoxiclav |
| Co-fluampicil |
| Co-trimoxazole |
| Demeclocycline |
| Doxycycline |
| Levofloxacin |
| Minocycline |
| Moxifloxacin |
| Nalidixic Acid |
| Nitrofurantoin |
| Norfloxacin |
| Ofloxacin |
| Oxytetracycline |
| Pivmecillinam |
| Tetracycline |
| Trimethoprim |

**Table S2-Antibiotic categories for multidrug resistance (MDR) definition for antibiotics with susceptibility data available**

| Antibiotic Class/Grouping | Included in consensus definitions table and study datasets | Additional Antibiotics in study susceptibility datasets |
|---|---|---|
| **Aminoglycosides** | Gentamicin<br>Tobramycin<br>Amikacin | HL kanamycin<br>Streptomycin HL |
| **Antipseudomonal penicillins + β-lactamase inhibitors** | Tazocin<br>Ceftolozane-tazobactam<br>Timentin<br>Ticarcillin | |
| **Non-extended spectrum cephalosporins; 1st and 2nd generation** | Cefuroxime | Cefalexin |
| **Extended spectrum cephalosporins; 3rd and 4th generation** | Ceftriaxone<br>Cefepime<br>Cefotaxime/MEN<br>Ceftazidime | Cefpodoxime |
| **Cephamycins** | Cefoxitin | |
| **Fluoroquinolones** | Ciprofloxacin | Ofloxacin<br>Levofloxacin |
| **Folate pathway inhibitors** | Trimethoprim<br>Trimethoprim-sulphamethoxazole | |
| **Monobactams** | Aztreonam | |
| **Penicillins** | | Amoxicillin<br>Pivmecillinam<br>Temocillin |
| **Penicillins+ β-lactamase inhibitors** | Amoxicillin-clavulanic acid | |
| **Tetracyclines** | Doxycycline<br>Minocycline<br>Tetracycline | |
| **Phosphonic Acids** | Fosfomycin | |
| **Glycylcyclines** | Tigecycline | |
| **Nitrofurantoin** | | Nitrofurantoin |
| **Phenicols** | Chloramphenicol | |
| **Carbapenems** | Ertapenem | |

**Figure S1- Numbers of A) urine specimens submitted, and B) *E. coli* urine isolates over time in NHS Tayside**

**A)**

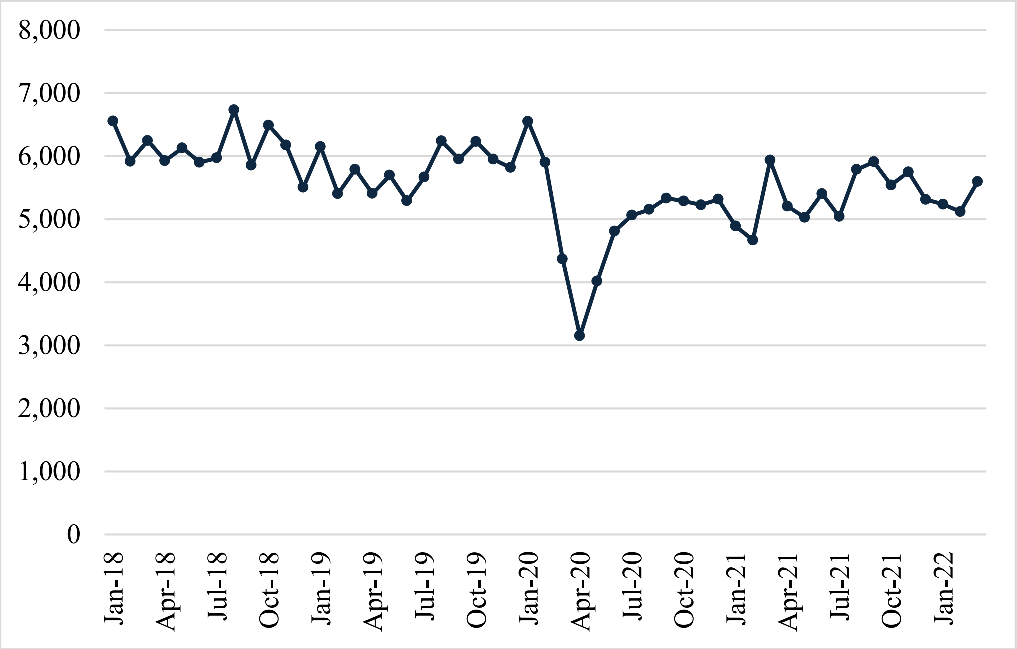


**B)**

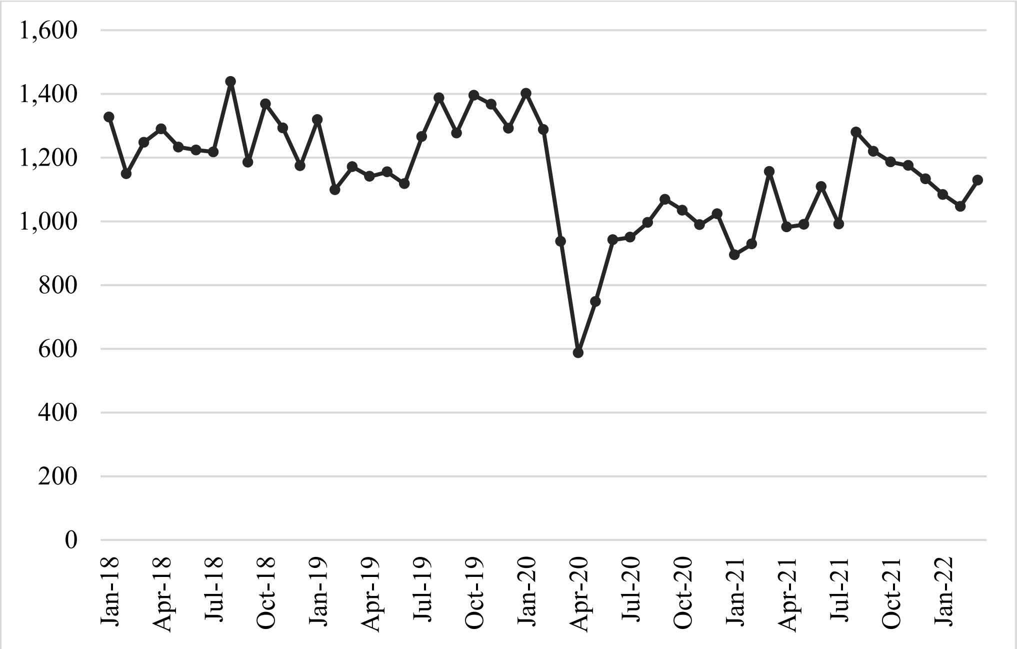

**Figure S2- Numbers of A) blood culture samples and B) *E. coli* blood culture isolates over time, NHS Tayside**

**A)**

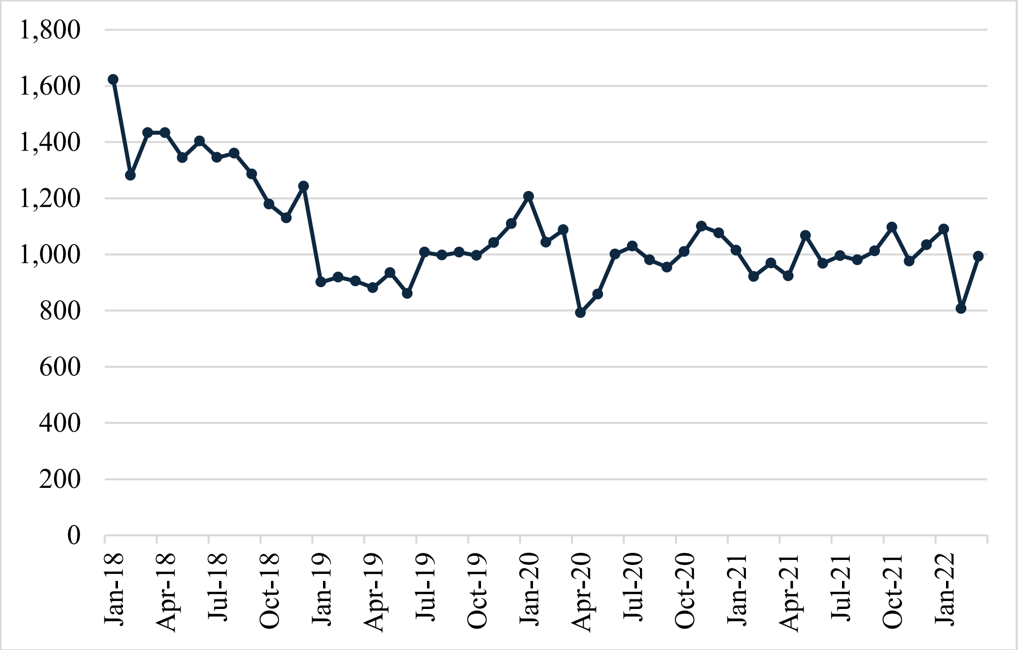


**B)**

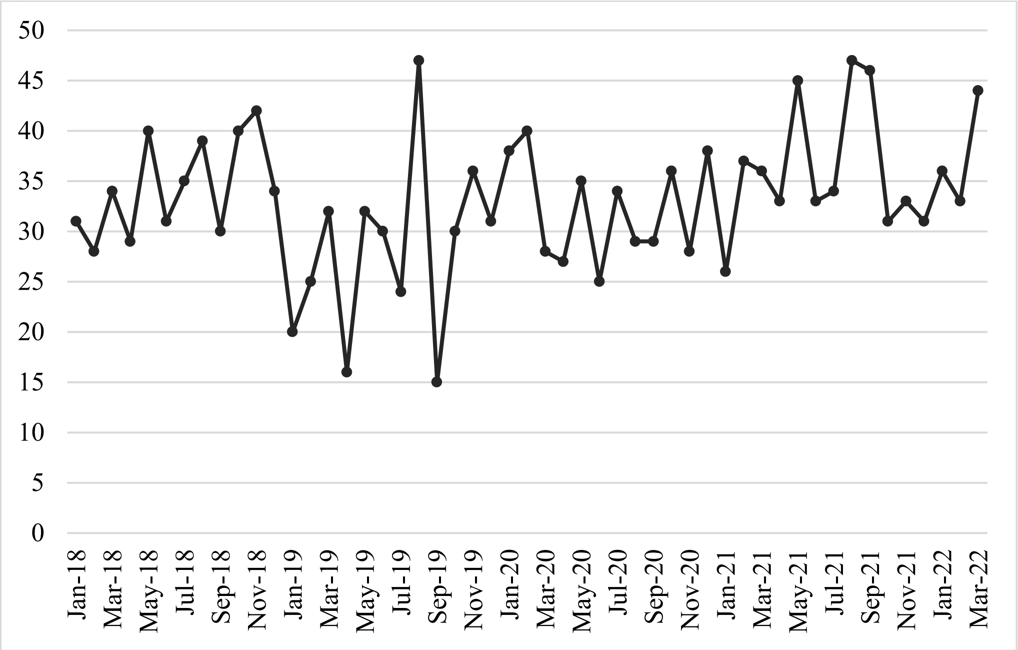

**Table S3- *E. coli* urine isolates: difference between actual modelled and projected modelled values of resistant at 3, 6, and 9 months after January 2020 joinpoint**

| **Month** | **Difference from Projected Counterfactual** | **Difference if Last Value Carried Forward** |
|---|---|---|
| Resistant (not MDR) | | |
| April 2020 (3 months) | +3.2% | +1.8% |
| July 2020 (6 months) | +4.2% | +2.1% |
| October 2020 (9 months) | +5.2% | +2.5% |
| MDR | | |
| April 2020 (3 months) | -0.1% | +0.2% |
| July 2020 (6 months) | -1.0% | -0.5% |
| October 2020 (9 months) | -1.9% | -1.3% |

**Table S4- *E. coli* blood culture isolates: difference between actual modelled and projected modelled values of resistant at 3, 6, and 9 months after January 2020 joinpoint**

| **Month** | **Difference from Projected Counterfactual** | **Difference if Last Value Carried Forward** |
|---|---|---|
| Resistant (not MDR) | | |
| April 2020 (3 months) | -0.5% | -2.3% |
| July 2020 (6 months) | +0.7% | -2.1% |
| October 2020 (9 months) | +2.0% | -2.0% |
| MDR | | |
| April 2020 (3 months) | -4.0% | -3.3% |
| July 2020 (6 months) | -5.3% | -3.6% |
| October 2020 (9 months) | -6.5% | -3.8% |

**Table S5- Autoregressive orders (p) and moving average orders (q) for interrupted time series analysis from testing for autocorrelation**

| **Data set** | **Autoregressive order (p)** | **Moving average order (q)** |
|---|---|---|
| *E. coli* Urine Isolates- Resistant (not MDR) | 0 | 1 |
| *E. coli* Urine Isolates- MDR | 0 | 1 |
| *E. coli* Blood Culture Isolates- Resistant (not MDR) | 1 | 2 |
| *E. coli* Blood Culture Isolates- MDR | 0 | 2 |
